# Downsizing of contact tracing during COVID-19 vaccine roll-out

**DOI:** 10.1101/2021.11.03.21265863

**Authors:** Maria M. Martignoni, Josh Renault, Joseph Baafi, Amy Hurford

## Abstract

Contact tracing is a key component of successful management of COVID-19. Contacts of infected individuals are asked to quarantine, which can significantly slow down (or prevent) community spread. Contact tracing is particularly effective when infections are detected quickly (e.g., through rapid testing), when contacts are traced with high probability, when the initial number of cases is low, and when social distancing and border restrictions are in place. However, the magnitude of the individual contribution of these factors in reducing epidemic spread and the impact of vaccination in determining contact tracing outputs is not fully understood. We present a delayed differential equation model to investigate how vaccine roll-out and the relaxation of social distancing requirements affect contact tracing practises. We provide an analytical criteria to determine the minimal contact tracing efficiency (defined as the the probability of identifying and quarantining contacts of symptomatic individuals) required to keep an outbreak under control, with respect to the contact rate and vaccination status of the population. Additionally, we consider how delays in outbreak detection and increased case importation rates affect the number of contacts to be traced daily. We show that in vaccinated communities a lower contact tracing efficiency is required to avoid uncontrolled epidemic spread, and delayed outbreak detection and relaxation of border restrictions do not lead to a significantly higher risk of overwhelming contact tracing. We find that investing in testing programs, rather than increasing the contact tracing capacity, has a larger impact in determining whether an outbreak will be controllable. This is because early detection activates contact tracing, which will slow, and eventually reverse exponential growth, while the contact tracing capacity is a threshold that will easily become overwhelmed if exponential growth is not curbed. Finally, we evaluate quarantine effectiveness during vaccine roll-out, by considering the proportion of people that will develop an infection while in isolation in relation to the vaccination status of the population and for different viral variants. We show that quarantine effectiveness decreases with increasing proportion of fully vaccinated individuals, and increases in the presence of more transmissible variants. These results suggest that a cost-effective approach during vaccine roll-out is to establish different quarantine rules for vaccinated and unvaccinated individuals, where rules should depend on viral transmissibility. Altogether, our study provides quantitative information for contact tracing downsizing during vaccine roll-out, to guide COVID-19 exit strategies.

## 1 Introduction

Contact tracing, in combination with quarantine, is a key component of the successful management of the COVID-19 pandemic. When contact tracing is in place, people with a confirmed infection provide information about individuals they have been in contact with during the previous days, who are in turn at risk of developing an infection. Identified contacts are traced and quarantined, and quick tracing can significantly slow down, or even prevent, epidemic spread, by quarantining infectious individuals before they become contagious. Efficient contact tracing may allow for partial relaxation of social distancing requirements and border restrictions, particularly during vaccine roll-out.

Different studies have focused on understanding the impact of contact tracing practices on COVID-19 outbreaks (Kretzschmar et al., 2020; Hellewell et al., 2020; Salathé et al., 2020; Fraser et al., 2004; Keeling et al., 2020; Tupper et al., 2020; Hu et al., 2021; Kucharski et al., 2020; Juneau et al., 2020; Gardner and Kilpatrick, 2021). Successful strategies involve quick detection of infectious cases (for example through testing) (Kretzschmar et al., 2020; Salathé et al., 2020; Fraser et al., 2004; Tupper et al., 2020; Hu et al., 2021; Gardner and Kilpatrick, 2021), a high probability that contacts are traced (Hellewell et al., 2020; Keeling et al., 2020; Hu et al., 2021; Gardner and Kilpatrick, 2021), a low number of initial cases when contact tracing is implemented (Hellewell et al., 2020), and, more generally, maintaining social distancing (Hellewell et al., 2020; Fraser et al., 2004; Keeling et al., 2020; Tupper et al., 2020), where different variants can affect viral transmissibility (Davies et al., 2021; Tegally et al., 2020). The implementation of border control measures can also impact contact tracing management (Aggarwal et al., 2021). Case importations contribute to epidemic spread when infection prevalence is low (Russell et al., 2021), and it is critical to consider how contact tracing practices should respond to the relaxation of border restrictions, in addition to the relaxation of social distancing. However, the link between case importations and the contact tracing efficiency needed to keep an outbreak under control has been little explored (Zhu et al., 2021). Finally, vaccination can clearly affect contact tracing management (Miller et al., 2020; Klinkenberg et al., 2006; Friston et al., 2021). During vaccine roll-out, we expect downsizing, or even dismantlement, of contact tracing to be possible. However, only few contact tracing models directly account for vaccine roll-out (Colomer et al., 2021; Ruebush et al., 2021). Additionally, vaccination may affect quarantine effectiveness, as isolation of vaccinated contacts, who have a significantly lower probability to get infected (Bernal et al., 2021; Nasreen et al., 2021), may lead to unnecessary costs related to isolation of healthy individuals (Wang et al., 2020).

While the factors increasing contact tracing efficiency during the COVID-19 outbreak have been identified, the magnitude of their individual contribution in reducing epidemic spread is not fully understood (Juneau et al., 2020). Most of the current approaches have been based on stochastic frameworks which, despite accounting for potential sources of uncertainty in disease transmission, limit the derivation of analytical expressions to quantitatively understand the interplay of different interventions during an outbreak (Fraser et al., 2004). Here, we aim to disentangle and quantify the impact of contact tracing practices during vaccine roll-out. We present a continuous time approach to investigate how the vaccination status of the population and the relaxation of social distancing requirements affect contact tracing programs. More specifically, we derive an analytical criteria for the minimal contact tracing efficiency needed to keep an outbreak under control, in relation to the vaccination status of a population and contact rate (section 3.1). Additionally, we quantify the impact of delays and case importations on the epidemics dynamics, to determine whether the contact tracing capacity should be adjusted during vaccine roll-out (section 3.2). We also consider how vaccine roll-out affects quarantine effectiveness, expressed as the proportion of people that will develop an infection while in quarantine (section 3.3).

Our study provides information about contact tracing downsizing during the relaxation of COVID-19 restrictions occurring concurrently to vaccine roll-out, and provides insights into the impact of contact tracing policies in different jurisdictions. The model presented here is general, and can be applied to different initial conditions, indicating differences in infection prevalence in a community. However, our focus will be on jurisdictions that have followed an elimination approach (Baker et al., 2020), where the infection is introduced in an initially virus-free community.

## 2 Model and Methods

To understand the impact of contract tracing on the epidemic during vaccine roll-out we extend the Susceptible-Exposed-Asymptomatic-Infected-Recovered (SEAIR) framework developed by Miller et al. (2020) to incorporate contact tracing (section 2.2), vaccination (section 2.3), delays in outbreak detection (section 2.4), and case importations (section 2.5). We will first describe the underlying SEAIR framework, and then explain how those additional components have been added to the model.

### 2.1 The underlying SEAIR model

If *c* is the daily number of contacts per person, and *α* is the probability of a positive transmission given a contact, it follows that the number of people infected each day is given by:

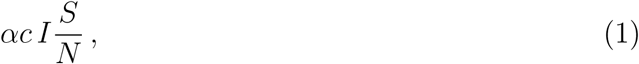

where variables *S, I* and *N* indicate the susceptible, infectious and total population. One of the factors that makes COVID-19 particularly difficult to trace is the abundance of asymptomatic individuals in the population, who are spreading the disease without awareness of their infectious status (He et al., 2021). To take asymptomatic cases into account, we differentiate infectious cases *I* into three subclasses, analogously to as done by Miller et al. (2020): asymptomatic *I*_*a*_ (i.e., infectious individuals who will never display COVID-19 symptoms), pre-symptomatic *I*_*p*_ (i.e., infectious individuals who have not yet developed symptoms), and symptomatic cases *I*_*c*_. The probability of an infection given a contact with an asymptomatic individual is lower than for symptomatic cases (Kronbichler et al., 2020), and we assume that symptomatic individuals have a reduced contact rate, as individuals feeling sick are less likely to interact with others (Miller et al., 2020). In the model, individuals in *I*_*a*_, *I*_*p*_ and *I*_*c*_ have a different infectivity, quantified by parameters *b*_*a*_, *b*_*p*_ and *b*_*c*_ respectively. Taking these factors into account, the number of people infected each day can be rewritten as:

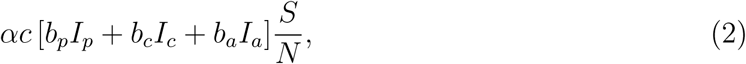

where *b*_*a*_ *< b*_*p*_ and *b*_*c*_ *< b*_*p*_.

Infected individuals enter the exposed class *E*, and after a latency period of average length 1*/δ*_*E*_ move either to the pre-symptomatic class *I*_*p*_, with probability *r*, or to the asymptomatic class *I*_*a*_, with probability (1*− r*). Once symptoms occur, pre-symptomatic individuals enter the symptomatic class. After the disease runs its course, individuals enter the recovered class *R*.

### 2.2 Modelling Contact Tracing

As only symptomatic individuals are contact traced, we write

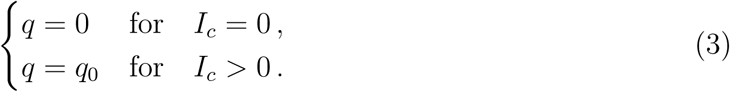

such that contract tracing is only activated when *I*_*c*_ *>* 0.

To identify contacts, we consider the interactions that individuals in *I*_*c*_ have had in the previous days with the susceptible population. Each symptomatic individual has entered class *I*_*c*_ by passing through the exposed class *E* and having entered the pre-symptomatic class with probability *r*. If we consider that the pre-symptomatic status lasts on average 2-3 days (Miller et al., 2020), and if we assume that symptomatic individuals become aware of their infectious status 1-2 days after symptoms onset, then *rδ*_*E*_*E*_(*t−*5)_ is number of people that entered *I*_*p*_ five days ago, and

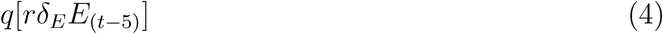

is equivalent to the number of people whose contacts are being traced today.

Individuals whose contacts are being traced are sent to quarantine, as well as susceptible, exposed, asymptomatic and pre-symptomatic individuals that have interacted with them in the past five days. The total number of identified contacts that contact traced individuals have had with susceptible people in the past five days can be expressed as

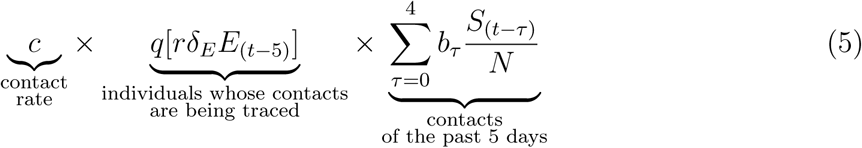

where *b*_*τ*_ = *b*_*p*_, *b*_*c*_ depending on whether the contact has been between susceptible and presymptomatic (meaning that at the time of the interaction the contact traced individual was still in *I*_*p*_, i.e., *b*_*τ*_ = *b*_*p*_ for *τ* = 2, 3, 4) or between susceptible and symptomatic cases (at the time of the interaction the contact traced individual had already entered *I*_*c*_, i.e., *b*_*τ*_ = *b*_*c*_ for *τ* = 0, 1).

As disease transmission given a contact occurs with probability *α*, a total of

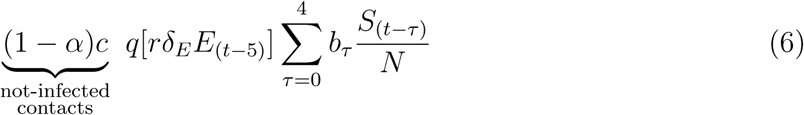

people per day will have interacted with the individual being contact traced without developing an infection. These people will move from the susceptible class *S* to class *S*_*q*_, which includes non-infectious individuals in isolation, and they will return to the susceptible class at the end of the quarantine period.

It follows that a total of

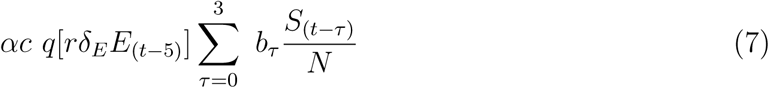

individuals will develop the infection and enter the exposed class. Individuals who have interacted with contact traced individuals five days before contact tracing began, will have already left the exposed class, and therefore a total of

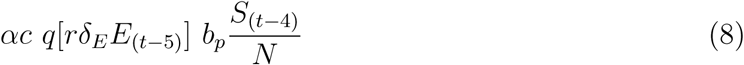

will be moved to quarantine after having already entered the pre-symptomatic class (with probability *r*) or the asymptomatic class (with probability (1 *− r*)). All contact traced individuals developing an infection move to the class *Q*, and then enter the recovered class *R* at the end of the quarantine period.

#### Model equations

The system of delay differential equations representing the model dynamics can be written as:

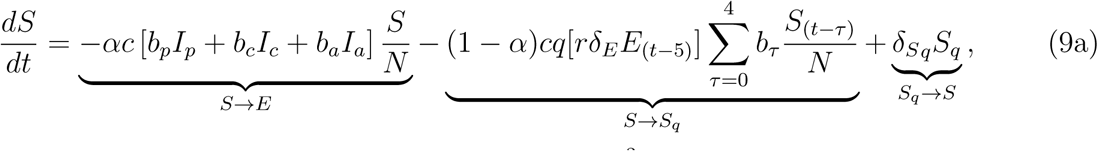

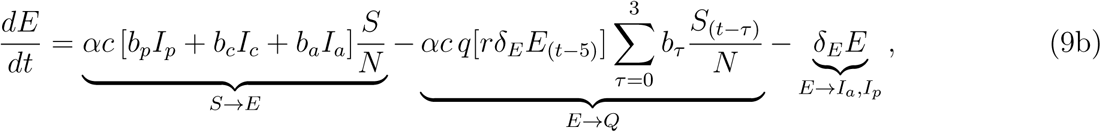

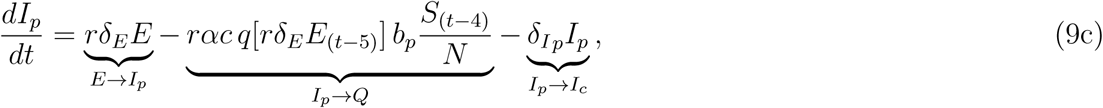

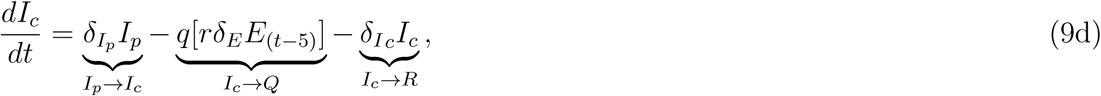

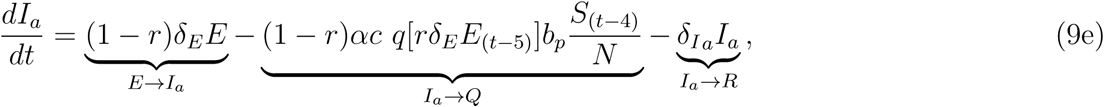

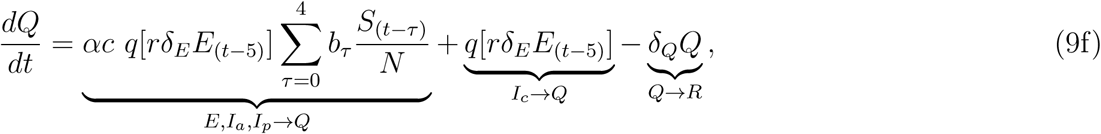

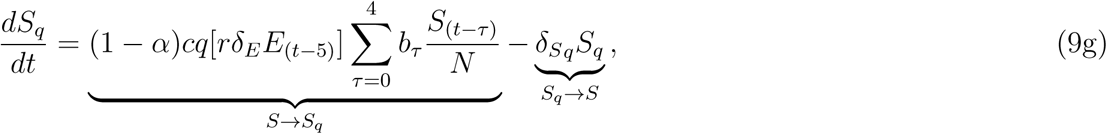

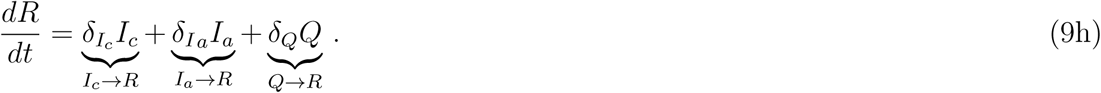

Note that the total number of individuals *N* remains constant over time. The flow diagram representation of Eq. (9) is presented in Fig. 1. Default parameters are given in Table 1. We will consider the case where the community is initially virus-free, and thus at time zero *S* = *S*_0_ (Eq. (10)), *E* = 1, and all other variables will be set to zero. Simulations will be performed with the dde23 solver of the software MATLAB R2018a.

**Table 1:** Brief description of the variables and parameters of the model given in Eq. (9), with corresponding default values or ranges considered for the simulations. Values in brackets correspond to the explored ranges.

| Variable/<br>parameter | Description | Value/<br>Range |
| --- | --- | --- |
| $S$ | Susceptible population | — |
| $E$ | Exposed population | — |
| $I_p$ | Pre-symptomatic infected population | — |
| $I_c$ | Symptomatic infected population | — |
| $I_a$ | Asymptomatic infected population | — |
| $Q$ | Infected population in quarantine | — |
| $S_q$ | Not-infected population in quarantine | — |
| $R$ | Recovered population | — |
| $N$ | Total population | 500,000 * |
| $\alpha$ | Probability of infection given a contact | 0.2 (0.1–0.5) * |
| $c$ | Contact rate | 5 (3–7) * |
| $b_p$ | Standard contact rate (pre-symptomatic) | 1 |
| $b_c$ | Reduction in contact rate (symptomatic) | 0.75 * |
| $b_a$ | Reduction in infectiousness (asymptomatic) | 0.5 (Davies et al., 2020) |
| $q$ | Contact tracing efficiency | 0.75 (0–1) |
| $r$ | Probability of becoming symptomatic given infected | 0.7 (He et al., 2021) |
| $I_{CT}^{max}$ | Contact tracing capacity | 250 (125, 500) * |
| $I_c^*$ | Symptomatic population when contact tracing is activated | 0 (0–9) |
| $\delta_E$ | Rate of people leaving state $E$ daily | 1/4 (Miller et al., 2020) |
| $\delta_{I_p}$ | Rate of people leaving state $I_p$ daily | 1/2.4 (Miller et al., 2020) |
| $\delta_{I_c}$ | Rate of people leaving state $I_c$ daily | 1/3.2 (Miller et al., 2020) |
| $\delta_{I_a}$ | Rate of people leaving state $I_a$ daily | 1/7 (Miller et al., 2020) |
| $\delta_Q$ | Rate of people leaving state $Q$ daily | 1/14 * |
| $\delta_{S_q}$ | Rate of people leaving state $S_q$ daily | 1/14 * |
| $p_v$ | Proportion of the population fully vaccinated | 0–1 |
| $\epsilon_v$ | Vaccine efficacy | 0.89 (Prunas et al., 2021) |
| $T$ | Total simulation time | 180 [days] |
| * Estimated parameters |  |  |

**Figure 1:**
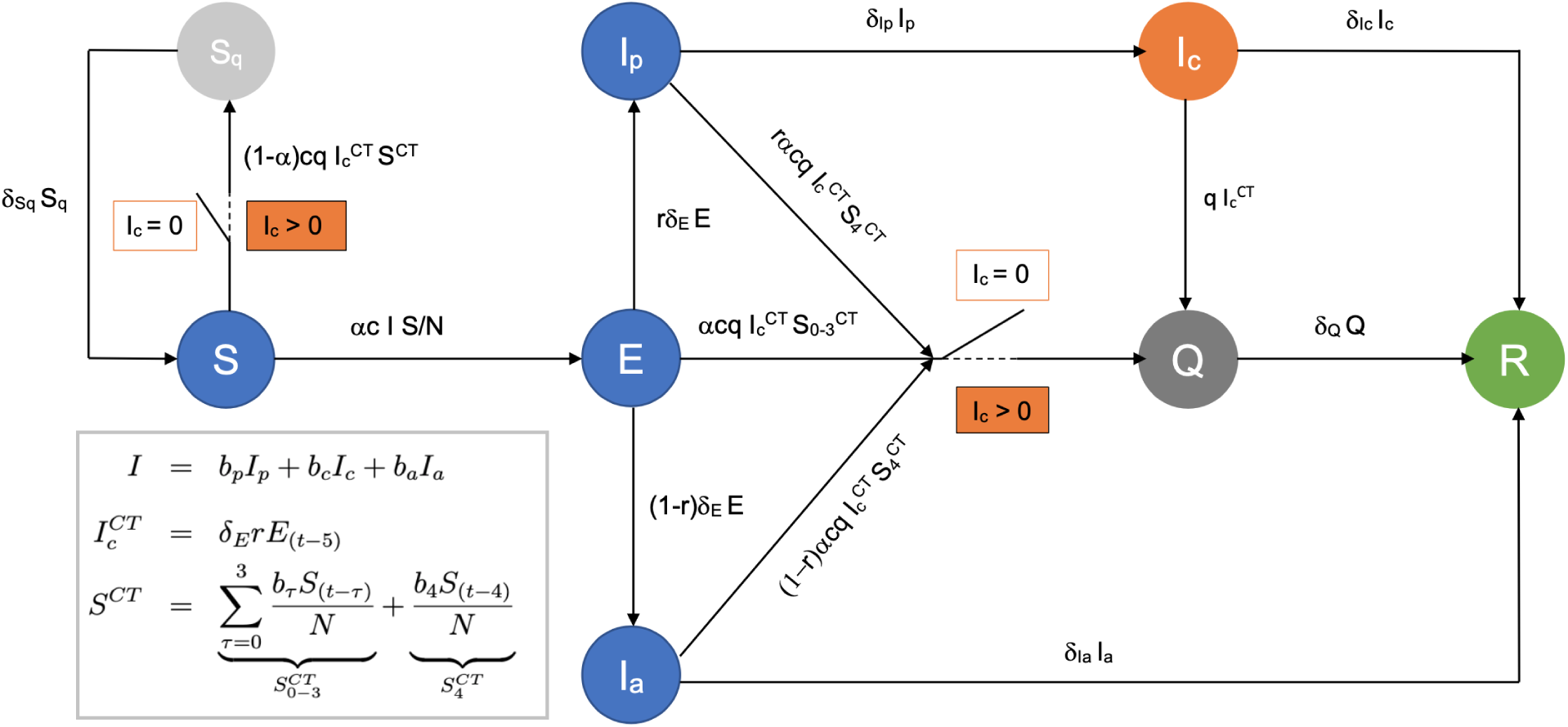
A schematic representation of the flow of individuals among different states in the population as described by equation (9). Susceptible individuals *S* can become exposed *E* after having interacted with an infected person, and successively become infectious pre-symptomatic *I*_*p*_, symptomatic *I*_*c*_ or asymptomatic *I*_*a*_. Individuals in states represented in blue are not aware of their infectious status and behave normally in the population. Individuals in *I*_*c*_, represented in orange, can become aware of their status and be contact traced. Once contact tracing is activated (for *I*_*c*_ *>* 0, see Eq. (3)) contacts can be moved to quarantine (states in grey), where contacts developing an infection will enter the *Q* class, and contacts not developing an infection will enter the *S*_*q*_ class. Recovered individuals enter the *R* status (in green). Delays in outbreak detection are modelled by assuming that contact tracing activates only when 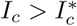, rather than when *I*_*c*_ *>* 0 (see section 2.4).

#### Contact tracing capacity

We assume that there is a limit in the contact tracing capacity, reached when the daily number of contacts to be traced (see Eq. (5) for *q* = 1) is higher than a certain value determined by 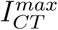. We will investigate the circumstances under which contact tracing is overwhelmed, and establish a criteria to determine the minimal contact tracing efficiency *q*\* needed to keep an outbreak under control. We understand a controllable outbreak as an outbreak where the daily number of contacts to be traced per day remains below the contact tracing capacity 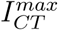 for *T* days. We will evaluate *q*\* with respect to the vaccination status of the population and the contact rate *c*. Default values of 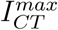 and *T* are provided in Table 1.

### 2.3 Modelling vaccination status

To simulate differences in the vaccination status of the population we assume that vaccination reduces the initial size of the susceptible population *S*_0_, where:

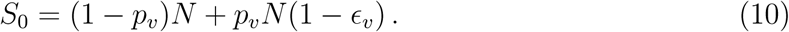

The parameter *p*_*v*_ represents the proportion of the susceptible population that is fully vaccinated. Vaccination reduces disease transmission with probability *ϵ*_*v*_. We assume that the vaccination status does not vary during the simulation period.

To account for contacts between vaccinated and contact traced individuals we compute the cumulative size of *S*_*q*_ (i.e., the number of people in quarantine that will never develop the infection) as follows:

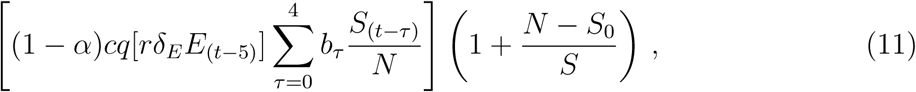

where the fraction (*N − S*_0_)*/S* accounts for vaccinated contacts in quarantine.

By comparing the cumulative number of infectious and non-infectious people in quarantine, we can gain information about quarantine effectiveness, intended as percentage of people quarantined that develop an infection (see supplementary information, Eq. (19)). We will investigate how quarantine effectiveness varies with the vaccination status of the population, and with the probability of infection given a contact (*α*), which can differ, for example, when considering different variants of the SARS-CoV2 virus.

### 2.4 Modelling delays in outbreak detection

Often outbreaks in a community are not promptly detected, and the disease can spread uncontrolled for several days before measures are introduced. We analyse this scenario by considering that contact tracing is activated only when a certain number of symptomatic cases 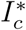 is found in the community. We modify the contact tracing efficiency *q*, defined in Eq. (3), as follows:

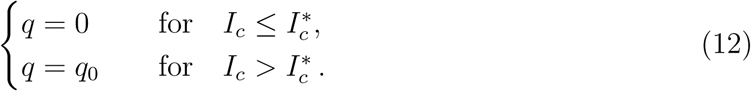

We will compute the daily number of contacts to be traced for different 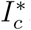, as a function of the vaccination status of the population and the contact rate, to understand how vaccine roll-out and social distancing impact the contact tracing capacity needed to keep an outbreak under control.

### 2.5 Modelling case importations

To evaluate the health risks following the relaxation of border restrictions during vaccine rollout, we investigate how case importations affect the daily number of contacts to be traced, for different vaccination status of the population and contact rates. When only one import is observed, we consider only one exposed case at the beginning of the simulation. For multiple imports *m*, we consider the *m* exposed cases as distributed evenly over *T* (where the first imports occurs at *t* = 0, the second import occurs at *t* = *T/m*, the third at *t* = 2*T/m*, and so on).

## 3 Results

### 3.1 Contact tracing efficiency and outbreak control

When the number of cases is low, we can derive an analytic expression for *q*\*, i.e., the minimal contact tracing efficiency needed to avoid a growth in the number of cases. If we assume that the number of infectious cases remains small over time, we can consider a substantially simplified version of the model of Eq. (9) consisting of a single differential equation in *I*, where *I* includes pre-symptomatic, symptomatic and asymptomatic infectious cases present in the time-dependent proportions 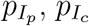 and 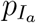 respectively. We write:

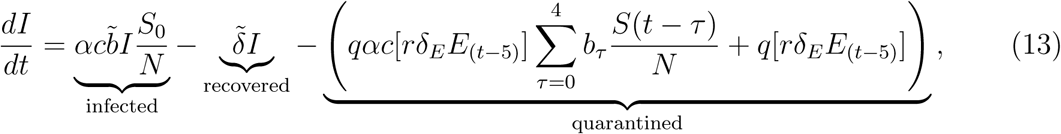

where *S*_0_ is the size of the susceptible population, which depends on vaccination (see Eq. (10)), parameter 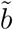 is defined as the weighted average of the contact rate (i.e., 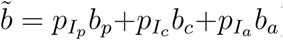), and parameter 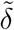 is the average time spent in the infectious state, and can be computed as:

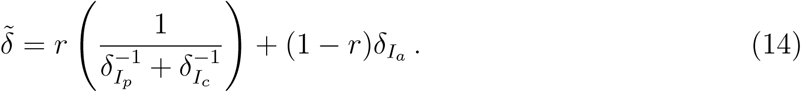

The factor *q*[*rδ*_*E*_*E*_(*t−*5)_] is the number of individuals contact tracing today, which, for *I* small, can be approximated as:

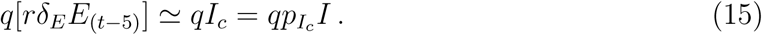

We obtain that *dI/dt <* 0 as long as:

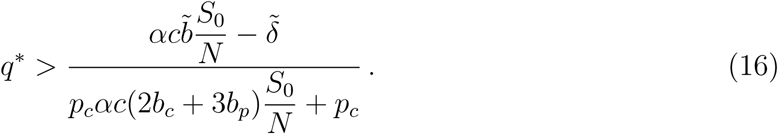

The minimal contact tracing efficiency needed to avoid disease spread therefore depends on: the vaccination status of the population, given by the ratio *S*_0_*/N* ; the contact rate *c*; the probability of infection given a contact *α*; the contact rates of pre-symptomatic, symptomatic and asymptomatic individuals *b*_*p*_, *b*_*c*_ and *b*_*a*_; the average length of the infectious status 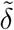;and the proportion of symptomatic individuals *p*_*c*_. A graphical representation of Eq. (16) is provided in the supplementary information, Fig. S.1.

When the contact rate *c* is high, Eq. (16) can be simplified as:

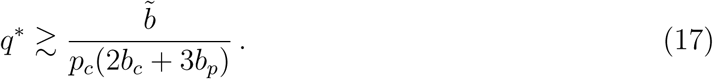

Given that 0 *≤ p*_*c*_ *≤* 1 and 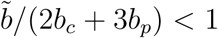, Eq. (17) indicates that there exists a minimal contact tracing efficiency 0 *< q*\* *<* 1 that can prevent infection spread, even in the absence of social distancing requirements, and independently from the vaccination status of the population. Thus, as long as *q > q*\*, contact tracing can be considered an effective sole intervention against COVID-19.

Our simulations confirm the relationships found in Eqs. (16) and (17). In Fig. 2 the minimal contact tracing efficiency *q* needed to avoid overwhelming contact tracing capacity is computed as a function of the vaccination status of the population and of the contact rate. We find that When *q* is large enough (i.e., *q* ≳ 0.5 in Fig. 2), overwhelming of contact tracing capacity does not occur, independently from the contact rate and vaccination status of the population (cfr. Eq. (17)). Thus, for *q* large, contact tracing alone is a sufficient measure to keep epidemic spread under control. For *q <* 0.5 we find that if vaccination rates are large enough, and/or if contact tracing is efficient enough, relaxation of social distancing requirements can occur. We see, for example, that the outbreak can be controlled without contact tracing if 55% of the population is vaccinated when the contact rate *c* = 3, or if 85% of the population is vaccinated when the contact rate *c* = 7. Alternately, with no vaccination, the outbreak can be controlled with contact tracing efficiency *q >* 0.4 when the contact rate is 3, and with *q >* 0.5 if the contact rate *c* = 7. Different variants (expressed as differences in the probability of infection given a contact *α*) can also affect the minimal contact tracing efficiency needed to avoid overwhelming contact tracing capacity, where higher contact tracing efficiency is needed to control outbreaks of more contagious variants.

**Figure 2:**
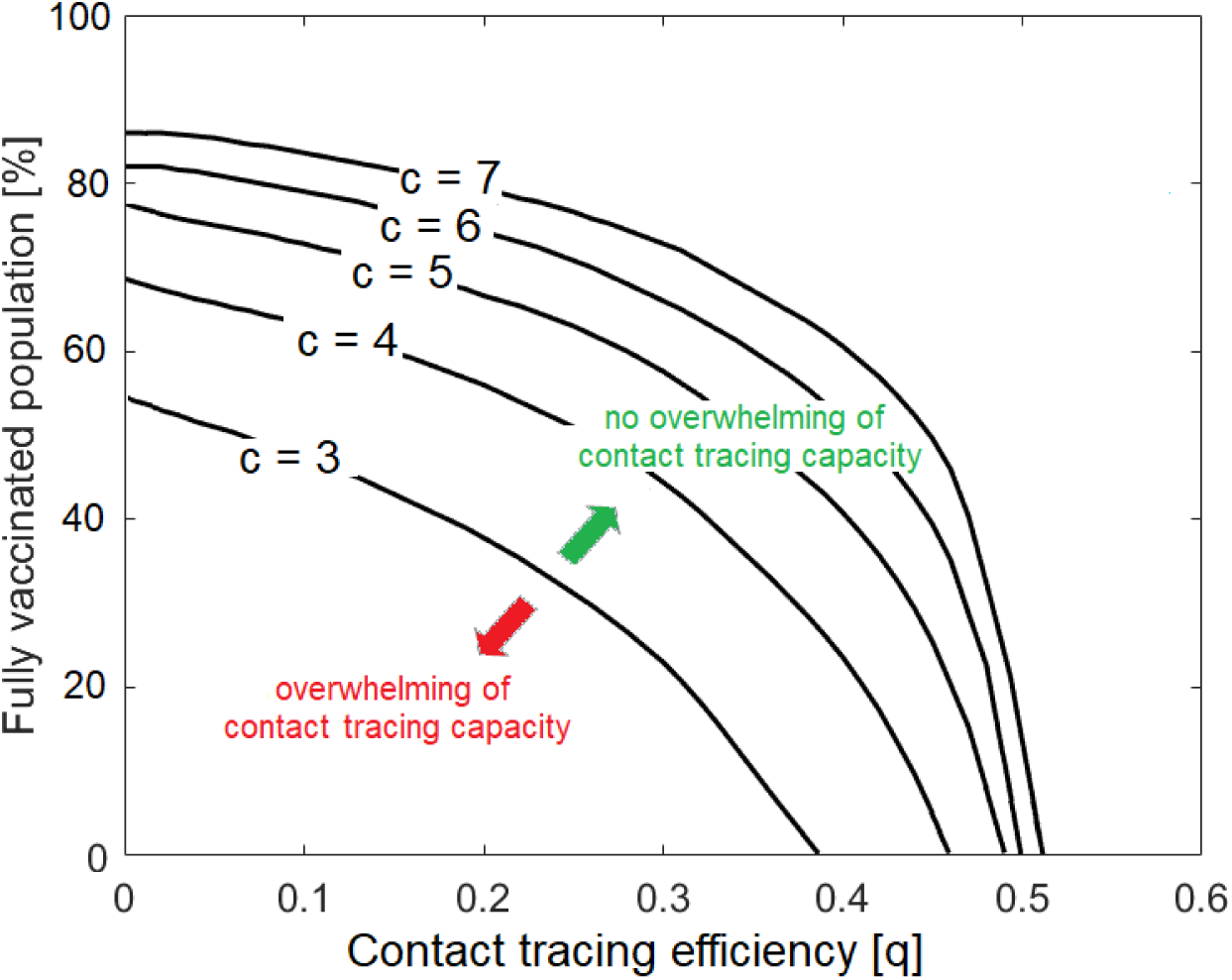
Minimal contact tracing efficiency needed to avoid overwhelming of contact tracing capacity, as a function of the vaccination status of the population and for different contact rates (black curves, with *c* = {3, 4, 5, 6, 7}), for 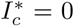. The area below each curve represents the parameter space for which contact tracing is overwhelmed, while the area above each curve represents the parameter space for which contact tracing is not overwhelmed. Default parameters used for the simulations are given in Table 1.

We found that the contact tracing capacity 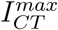 does not significantly affect the results (Fig. S.2). In a parameter space where the outbreak can be controlled, disease spread, and consequently overwhelming of the contact tracing capacity, will not occur. If the outbreak can not be controlled, the number of cases will grow nearly exponentially and exceed the contact tracing capacity 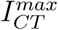, where doubling or halving 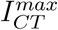 will not significantly affect the results. Therefore, the minimal contact tracing efficiency needed to avoid a growth in the number of cases (*q*\*, Eq. (16)), and the minimal contact tracing efficiency needed to avoid overwhelming contact tracing (obtained numerically) appear to be very similar quantities (cfr. Figs 2, S.1 and S.2). Efficiency, in terms of quick detection of symptomatic cases and identification and quarantining of their contacts, and not contact tracing capacity, is therefore the most important determinant of whether an outbreak will be controlled or not. Highly efficient contact tracing should keep the number of infections, and thus the number of contact tracing individuals, well below capacity.

### 3.2 Delayed detection and case importations

We consider the situation in which contact tracing is efficient enough to control the outbreak (i.e., *q* = 0.75, cfr. Fig. 2) and investigate the impact of delayed detection on contact tracing capacity, where delays are modeled as an increase in 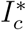 (see Eq. (12)), i.e., the number of symptomatic cases present in the community when contact tracing is activated. In Fig. 3a-c we see that if a delay in detection is experienced, the number of cases to be traced per day increases, particularly when the vaccination status of the population is low. An increase in the daily number of cases can lead to a non-controllable outbreak, even when the contact tracing efficiency is high. For example, we see that if the outbreak is detected only when already 6 symptomatic cases are present (see. Fig. 3b), in the absence of vaccination and with a contact rate of 7, the number of contacts to be traced is around 400 a day. Vaccination however, minimizes the impact of delays, and helps to maintain contact tracing within capacity, even when the contact rate is high. For example, if 60% of the population is vaccinated and the outbreak is detected when 6 symptomatic cases are already in the community, the maximum number of contacts to trace with a high contact rate of 7 per day is around 50, and thus more easily manageable (see Fig. 3b).

**Figure 3:**
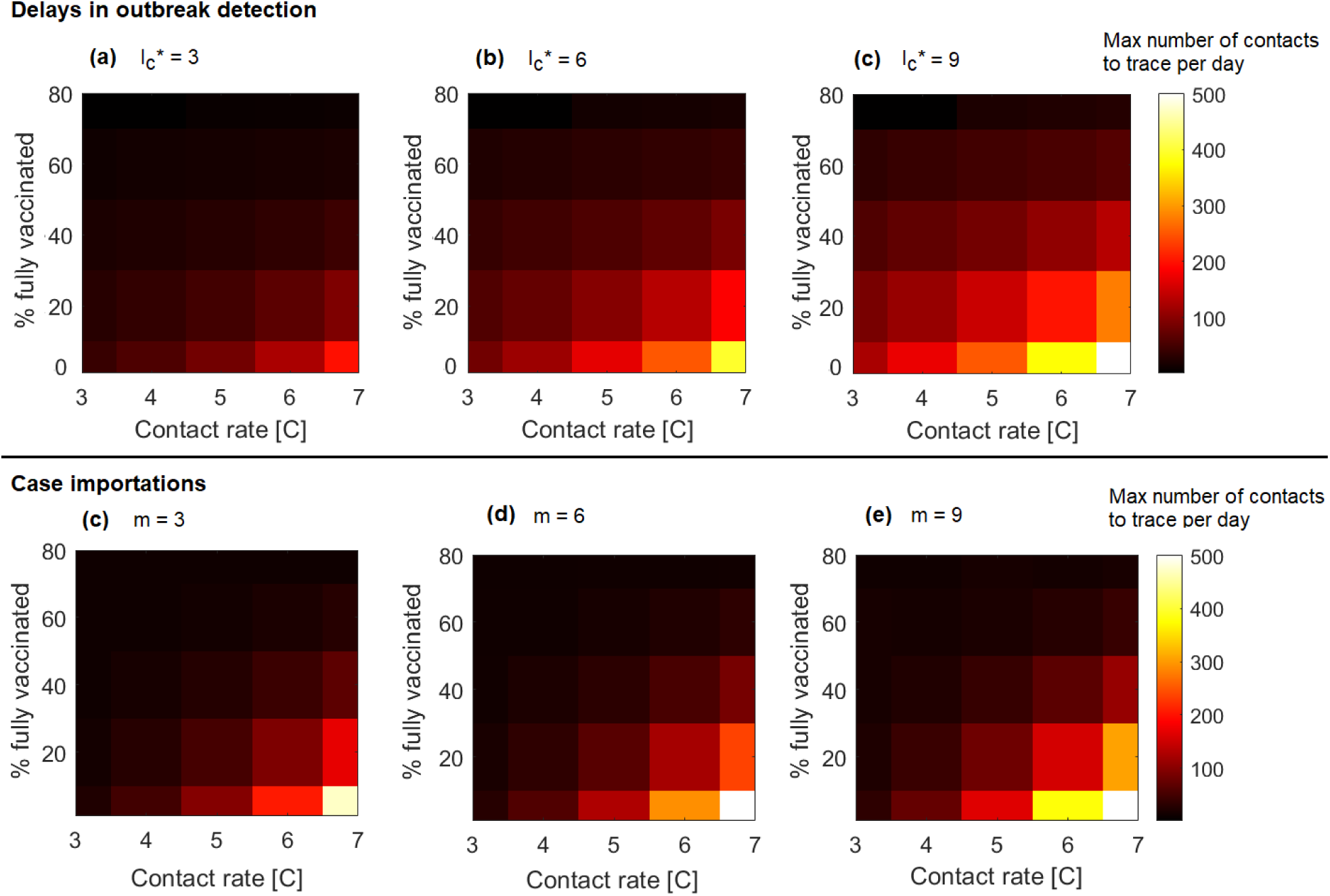
Maximal number of contacts to trace per day as a function of the percentage of fully vaccinated individuals in the population and the contact rate. The number of symptomatic individuals already present in the community when contact tracing is activated 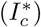 is progressively increased from (a) 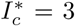, to (b)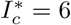, to 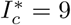. In figures (d)-(f) 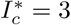 for all simulations, and the number of imported cases *m* over the time interval considered (i.e., *T* = 180 days) varies, with (d) *m* = 3, (e) *m* = 6, and (f) *m* = 9. The contact tracing efficiency is kept constant at *q* = 0.75.

Case importations can also lead to overwhelming contact tracing capacity (Fig. 3d-e), however vaccination can strongly reduce this risk. Indeed, if for example a number of 6 imports are experienced in 180 days, the number of contacts to trace per day might reach 500 in the absence of vaccination, while it remains lower than 40 when 60% of the population is vaccinated (see Fig. 3e). Note however that when a region experiences multiple imports, each import incorporates a risk for delayed detection, leading to an increased risk of exceeding capacity (as seen in Fig. 3a-c). Swift detection of infected imported cases is therefore important to make sure that the contact tracing capacity is not overwhelmed. Additionally, in the simulations we assumed imports to be evenly distributed over the time interval considered. However, imports might occur simultaneously, which could increase the risk of overwhelming of contact tracing capacity.

### 3.3 Quarantine effectiveness

We look at how quarantine effectiveness, intended as the proportion of quarantined individuals that develop an infection, is affected by the vaccination status of the population and by disease infectiousness (i.e., the probability of infection given a contact *α*) (Fig. 4). Quarantine effectiveness decreases when the proportion of fully vaccinated people in the population is high. Additionally, quarantine effectiveness increases when *α* is high, meaning that the percentage of quarantined individuals that develops an infection is higher in the presence of more contagious variants. For example, we obtain that for *α* = 0.2, about 25% of the quarantined individuals will develop an infection in an unvaccinated population. With 75% of the population being fully vaccinated, only 10% of the quarantined individuals will develop an infection, thereby decreasing quarantine effectiveness by more than half. Note that quarantine effectiveness does not depend on the contact tracing efficiency *q* (see supplementary information, Eq. (19)).

**Figure 4:**
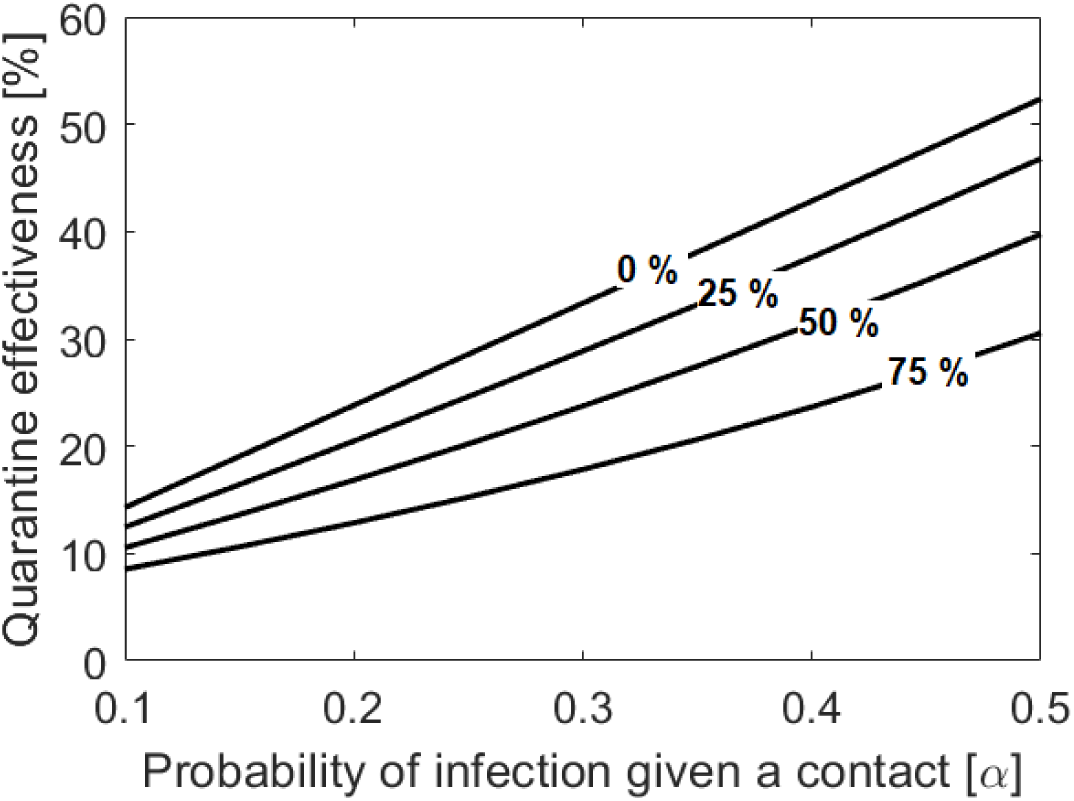
Quarantine effectiveness, defined as the percentage of quarantined individuals that develop an infection, as a function of the probability of infection given a contact (*α*). Different curves represent different vaccination status of the population, where we consider that 0%, 25%, 50% or 75% of the population has been fully vaccinated.

## 4 Discussion

Previous work has disputed whether contact tracing can be used as sole intervention to control outbreaks (Tupper et al., 2020; Davis et al., 2021; Juneau et al., 2020; Ferretti et al., 2020). Ferretti et al. (2020) found that epidemic control through contact tracing could be achieved through the immediate notification and isolation of at least 70% of infectious cases, while three or more days delay in case notification would not allow for epidemic control. Analogously, we show that, when highly efficient, contact tracing alone can be considered an effective control measure even in the absence of vaccination or social distancing. For example, for an average contact rate of 7 individuals per day, contact tracing can be an effective sole intervention as long as more than 50% of the contacts of symptomatic individuals are quarantined within 1 or 2 days from symptoms onset. However, delays in detection and relaxation of border control measures can cause the number of contacts to be traced in a day to exceed the contact tracing capacity. Similarly, other studies found that testing at first symptom is a necessary prerequisite for efficient tracing (Davis et al., 2021; Gardner and Kilpatrick, 2021; Kretzschmar et al., 2020; Kucharski et al., 2020; Hellewell et al., 2020), and that a higher contact tracing efficiency is needed to keep an outbreak under control when the number of initial cases is large (Hellewell et al., 2020; Dhillon and Srikrishna, 2018). These findings emphasize the importance of testing at first symptoms, as well as testing new arrivals, to avoid overwhelming contact tracing capacity.

We find that investing in fast detection, for example via testing programs, rather than increasing the contact tracing capacity, has a larger impact in determining whether an outbreak will be controllable. Strong testing programs to ensure the quick detection of new community outbreaks, in combination with efficient identification and isolation of contacts, ensures slow epidemic spread, where the number of daily contacts to be traced remains low for the whole duration of the outbreak. Should slow detection cause uncontrolled epidemic spread, we expect overwhelming of contact tracing capacity to occur even when the maximum daily number of tracing contacts is large, owing to exponential growth of the outbreak.

Vaccination has the double impact of reducing the contact tracing efficiency required to keep an outbreak under control, and minimizing the impact of delays and case importations. Indeed, in vaccinated communities, a lower contact tracing efficiency is required to avoid overwhelming contact tracing capacity. For example, with 70% of the population fully vaccinated, a contact tracing efficiency of 40% is enough to keep an outbreak under control, even with a high average contact rate of 7 individuals per day. Additionally, predictions show that the maximum number of contacts to be traced per day is drastically reduced when epidemic spread occurs in vaccinated populations, where delays in detection or increase in the number of imported cases in vaccinated populations do not lead to a significant risk of overwhelming contact tracing capacity. These findings suggest possible downsizing of contact tracing practices during vaccine roll-out, even when vaccination occurs in conjunction with the relaxation of social distancing and border restrictions. As vaccination rates are distributed heterogeneously in the population, contact tracing downsizing, rather than dismantlement, should be considered, especially as contact tracing remains an important measure to reduce or avoid community spread in low vaccination settings, such as schools for young children that may not be vaccine eligible.

Efficient tracing can be affected at many stages of the contact tracing process. Individuals may delay getting tested, and positive results may take days to be confirmed (Lewis, 2020). Additionally, contacts may not be easily identified or contacted, and they may not adhere to isolation requirements (Lewis, 2020; Davis et al., 2021; Gardner and Kilpatrick, 2021). Generally, higher efficiency can be achieved in regions characterised by social cohesiveness, such as small jurisdictions with interconnected populations, where infected individuals might be known and a high proportion of contacts is likely to be reached (WHO, 2021; Lash et al., 2021). In denser populations, contact identification may be an arduous task, where manual contact tracing might be impractical and electronic contact tracing, for example through mobile apps, has often raised privacy concerns (Zastrow, 2020; Cho et al., 2020). Thus, while contact tracing might be an effective sole intervention in rural areas, failure might be observed in larger or more densely populated regions, which emphasizes the potential need for different policy decisions in small and large jurisdictions.

In our model, we assume contacts of symptomatic individuals to be isolated within 1 - 2 days, and we do not explicitly take into account possible delays from testing of symptomatic individuals to quarantining of their contacts. Additionally, we assume that individuals in quarantine do not transmit the disease, while this might often not be the case. Possible extensions of the model presented here include delays in the identification of contacts, and poor community adherence to quarantine rules (Davis et al., 2021).

Finally, we show that quarantine effectiveness is low in a vaccinated population, as a large proportion of quarantined contacts will not develop an infection. These findings suggest that a cost-effective approach during vaccine roll-out is to establish different quarantine rules for vaccinated and unvaccinated individuals, as has indeed been done in several jurisdictions (for Disease Control et al., 2021). Rules should be evaluated with respect to the presence of more transmissible viral variants, which can increase the probability of infection given a contact for unvaccinated as well as for vaccinated individuals (Davies et al., 2021; Tegally et al., 2020). Future modelling efforts should explicitly consider the risk of non-quarantining vaccinated individuals, in the presence of different viral variants.

## Data Availability

All data produced in the present work are contained in the manuscript

## 5 Acknowledgement

JR is supported by a National Sciences and Engineering Research Council of Canada (NSERC) Undergraduate Student Research Award (USRA). AH acknowledges financial support from an NSERC Discovery Grant, RGPIN 2014-05413. MM and AH are supported by Canadian Network for Modelling Infectious Diseases - Réseau canadien de modélisation des maladies infectieuses (CANMOD) and the Department of Health and Community Services, Government of Newfoundland and Labrador. AH acknowledges further support from the NSERC Emerging Infectious Disease Modelling Consortium. AH and JB are supported by the Atlantic Association for Research in the Mathematical Sciences and the New Brunswick Health Research Foundation.

## Supplementary information

### Contact tracing efficiency and controllable outbreaks

**Figure S.1:**
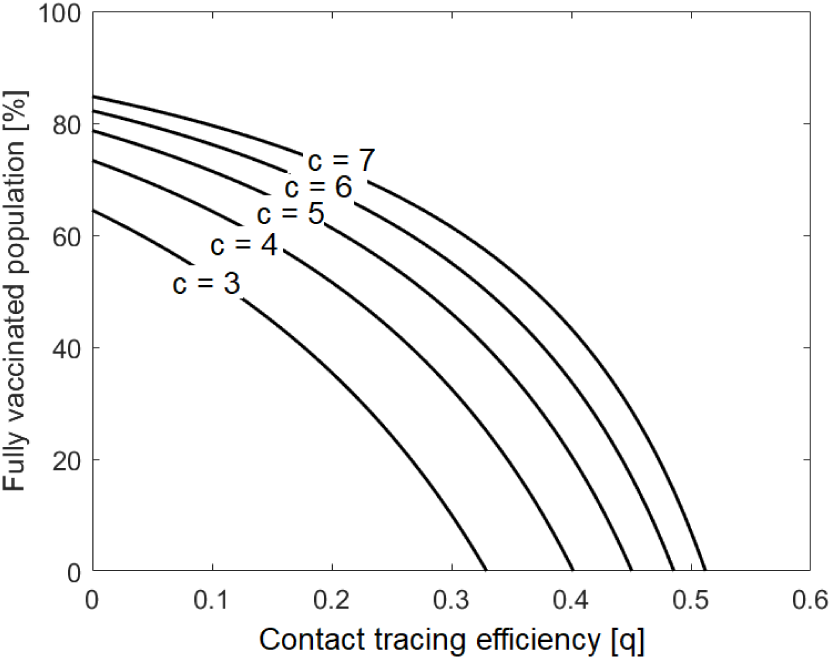
Analytical approximation of the minimal contact tracing efficiency *q*\* needed to avoid disease spread, as a function of the vaccination status and for different contact rates. Lines represent equality in Eq. (16), for the contact rates *c* = {3, 4, 5, 6, 7}. Default parameters are given in Table 1.

**Figure S.2:**
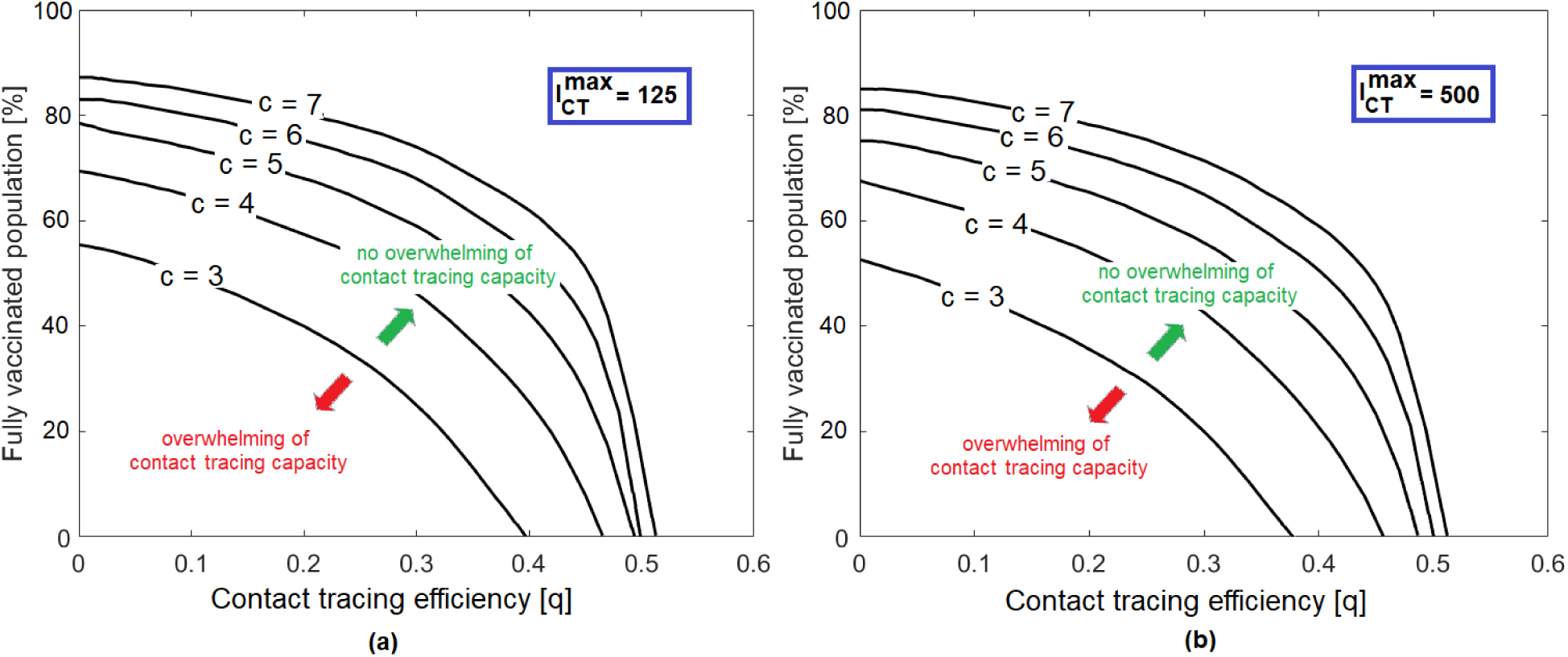
Minimal contact tracing efficiency needed to avoid overwhelming of contact tracing capacity, as a function of the vaccination status and for different contact rates (black curves, with *c* = {3, 4, 5, 6, 7 }) for (a) a higher contact tracing capacity 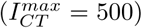 and (b) a lower contact tracing capacity 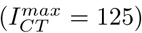. The area below each curve represents the parameter space for which contact tracing is overwhelmed, while the area above each curve represents the parameter space for which contact tracing is not overwhelmed. Parameter 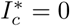 for all simulations, other default parameters are given in Table 1. We see that, as explained in section 3.1, differences in the contact tracing capacity 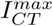 do not significantly affect the results (cfr. Eq. (16)).

### Quarantine effectiveness

We calculate the quarantine effectiveness as the number of people in quarantine that develop an infection (which corresponds to the cumulative number of individuals entering class *Q*, derived from Eq. (9f)) divided by the total number of people in quarantine (i.e., the sum of the cumulative number of individuals entering class *Q* and the cumulative number of individuals entering class *S*_*q*_, given in Eq. (11)). Mathematically, we obtain the following expression for quarantine effectiveness *Q*_*eff*_ :

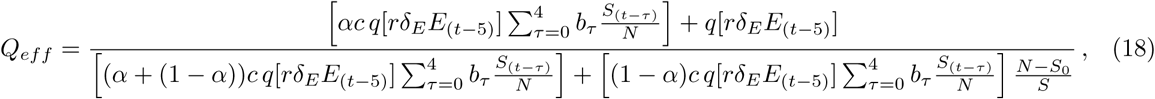

which can be simplified as

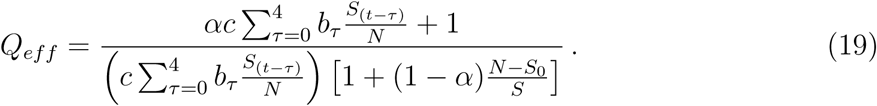

Quarantine effectiveness depends therefore on the probability of infection given a contact (*α*) and on the vaccination status of the population, expressed through the number of susceptible individuals *S*_0_. Note that quarantine effectiveness does not depend on the contact tracing efficiency *q*.

